# Combining Tumor Segmentation Masks with PET/CT Images and Clinical Data in a Deep Learning Framework for Improved Prognostic Prediction in Head and Neck Squamous Cell Carcinoma

**DOI:** 10.1101/2021.10.14.21264958

**Authors:** Kareem A. Wahid, Renjie He, Cem Dede, Abdallah Mohamed, Moamen Abobakr Abdelaal, Lisanne V. van Dijk, Clifton D. Fuller, Mohamed A. Naser

**Affiliations:** Department of Radiation Oncology, The University of Texas MD Anderson Cancer, Houston, Texas 77030, USA

**Keywords:** PET, CT, Head and Neck Cancer, Oropharyngeal Cancer, Deep Learning, Progression-Free Survival, Outcome Prediction Model, Segmentation Mask

## Abstract

PET/CT images provide a rich data source for clinical prediction models in head and neck squamous cell carcinoma (HNSCC). Deep learning models often use images in an end-to-end fashion with clinical data or no additional input for predictions. However, in the context of HNSCC, the tumor region of interest may be an informative prior in the generation of improved prediction performance. In this study, we utilize a deep learning framework based on a DenseNet architecture to combine PET images, CT images, primary tumor segmentation masks, and clinical data as separate channels to predict progression-free survival (PFS) in days for HNSCC patients. Through internal validation (10-fold cross-validation) based on a large set of training data provided by the 2021 HECKTOR Challenge, we achieve a mean C-index of 0.855 ± 0.060 and 0.650 ± 0.074 when observed events are and are not included in the C-index calculation, respectively. Ensemble approaches applied to cross-validation folds yield C-index values up to 0.698 in the independent test set (external validation). Importantly, the value of the added segmentation mask is underscored in both internal and external validation by an improvement of the C-index when compared to models that do not utilize the segmentation mask. These promising results highlight the utility of including segmentation masks as additional input channels in deep learning pipelines for clinical outcome prediction in HNSCC.

## 1 Introduction

Deep learning has been studied extensively for medical image segmentation and clinical outcome prediction in head and neck squamous cell carcinoma (HNSCC) [1]. While utilizing medical images in an end-to-end framework with no additional data streams has become commonplace, previous studies have noted the importance of adding regions of interest in a deep learning workflow to improve predictive performance [2, 3]. These regions of interest may help deep learning models localize to areas that harbor more relevant information for the downstream prediction task of interest. This may be particularly salient for HNSCC patients, where the prognostic status is often informed by the re-appearance of tumor volumes in or near the original region of interest (recurrence/treatment failure) [4]. Moreover, PET/CT provides a rich source of information for the tumor region of interest in HNSCC [5] that may be combined with deep learning models to improve prognostic prediction, such as for progression-free survival (PFS). Therefore, the development of PET/CT-based deep learning approaches that can effectively combine previously segmented tumor regions of interest with existing architectures is an important component in exploring novel and effective HNSCC outcome prediction models.

In this study, we develop and evaluate a deep learning model based on the DenseNet architecture that combines PET/CT images, primary tumor segmentation masks, and clinical data to predict PFS in HNSCC patients provided by the 2021 HECKTOR Challenge. By combining these various information streams, we demonstrate reasonable performance on internal and external validation sets.

## 2 Methods

We developed a deep learning model for PFS prediction of HNSCC patients using co-registered ^18^F-FDGPET and CT imaging data, ground truth primary tumor segmentation masks, and associated clinical data. The censoring status and time-to-event between PET/CT scan and event, imaging data, segmentation masks, and clinical data were used to train the model. The performance of the trained model for predicting PFS was validated using a 10-fold cross-validation approach. An ensemble model based on the predictions from the 10-fold cross-validation models was packaged into a Docker container to evaluate models on unseen testing data. Most methodological details of our study have been previously outlined in a parallel study on Task 2 of the HECKTOR Challenge (PFS prediction without using segmentation mask information). Abbreviated methodology salient to the use of segmentation masks in our models is as follows.

Data from 224 HNSCC patients from multiple institutions was provided in the 2021 HECKTOR Challenge [6, 7] training set. Data for these patients included co-registered ^18^F-FDG PET and CT scans, clinical data (Center ID, Gender, Age, TNM edition, chemotherapy status, TNM group, T-stage, N-stage, and M-stage), and ground truth manual segmentations of primary tumors derived from clinical experts. Images and masks were cropped to a bounding box of 144×144×144 mm^3^ and resampled to a resolution of 1 mm. The CT intensities were truncated in the range of [-200, 200] and then normalized to a [-1, 1] scale, while PET intensities were normalized with a z-score. Clinical data were mapped to ordinal categorical variables and min-max rescaled. The clinical data was reshaped into a 144×144×144 volume by concatenated clinical variables repeatedly to act as a volumetric input to the deep learning model.

A deep learning convolutional neural network model based on the DenseNet121 [8] architecture included in the MONAI Python package [9] was used for the analysis (**Fig. 1**). Processed PET/CT, tumor mask, and clinical data volumes were used as separate input channels to the model. We used data augmentation by MONAI [9] during training which was applied to the PET/CT and tumor mask images. The augmentation included random horizontal flips of 50% and random affine transformations with an axial rotation range of 12 degrees and a scale range of 10%. We used a batch size of 2 patients’ images, masks, and clinical data. The model was trained for 800 iterations with a learning rate of 2×10^−4^ for iterations 0 to 300, 1×10^−4^ for iterations 301 to 600, and 5×10^−5^ for iterations 601 to 800. We used an Adam optimizer and a negative log-likelihood loss function.

**Fig. 1.**
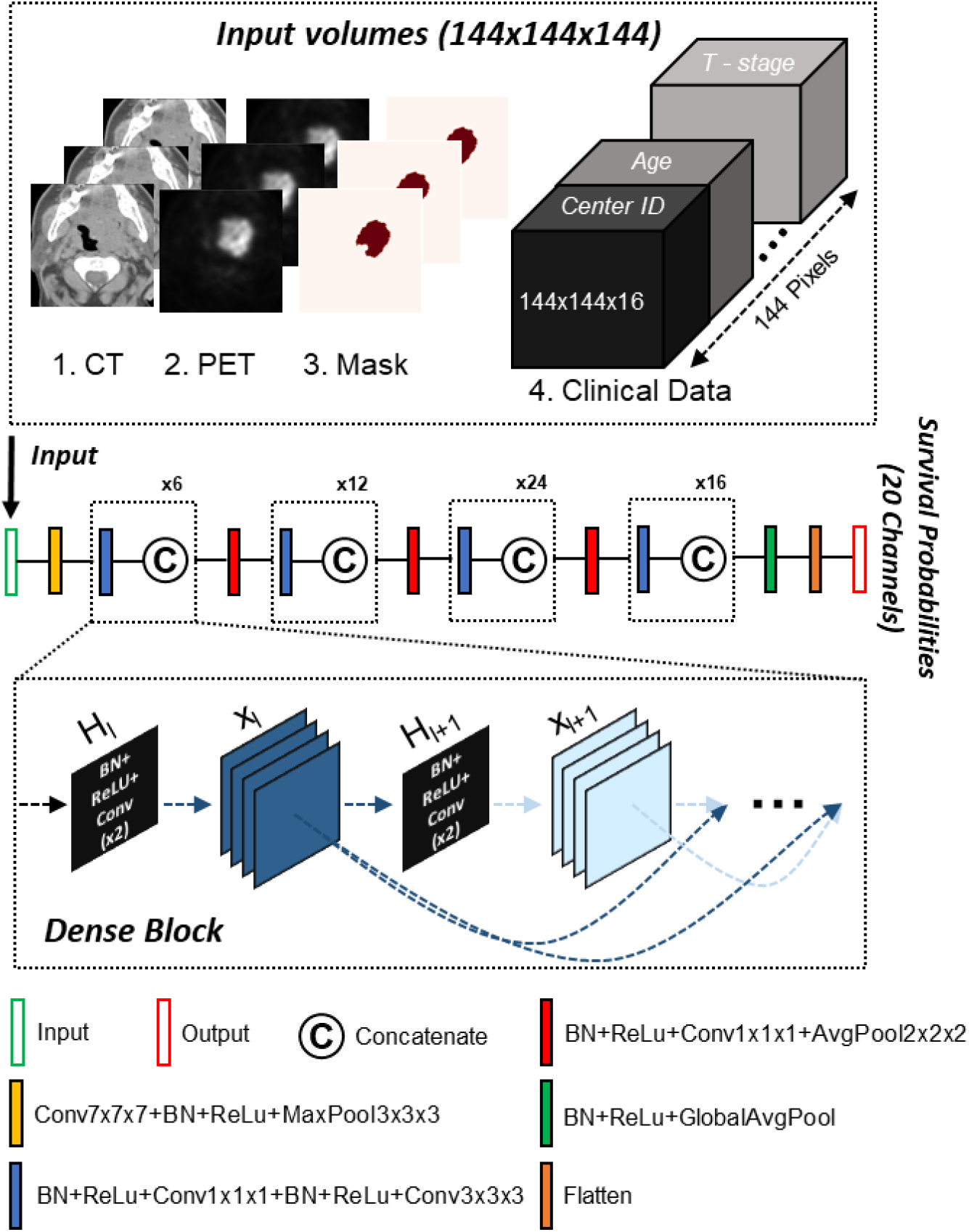
Schematic of the Densenet121 architecture used for the prediction model and the four-channel input volumetric images representing CT, PET, tumor mask, and clinical data. Ground truth tumor segmentation masks for model training were provided by the 2021 HECKTOR Challenge and accessed via a Docker framework for test set evaluation. The number of repeated dense blocks of (6, 12, 24, and 16) are given above each group of blocks.

As defined on the HECKTOR Challenge website, progression was defined based on RECIST criteria: either a size increase of known lesions (change of T and or N), or appearance of new lesions (change of N and/or M), where disease-specific death was also considered a progression event. To model PFS we divided the total time interval of 9 years, which covers all values reported for PFS in the training data set, into 20 discrete intervals of 164.25 days, representing the final 20 output channels of the network. The conditional probabilities of surviving in these intervals were obtained by applying a sigmoid function on the 20 outputs channels of the network. We estimated the final PFS from the model predicted conditional probabilities by obtaining the summation of the cumulative probability of surviving each time interval times the duration of the time interval of 164.25 days. We used a 10-fold cross-validation approach to train and evaluate the model. We assessed the performance of each separate cross-validation model using the concordance index (C-index) from the lifelines Python package [10]. We estimated the mean C-index by averaging all the C-index values obtained from each fold. It is possible to measure the C-index by ignoring the events observed; therefore, as an alternative metric, we also measured this modified C-index in reporting results.

For the independent test set on the HECKTOR submission portal, we implemented two different model ensembling approaches to estimate the PFS from cross-validation results: a simple average across models (AVERAGE), and a consensus from cumulative survival probability derived from mean conditional probability survival vectors (CONSENSUS). We packaged these models into Docker images [11] that act as templates to apply the models to unseen test data. Each Docker image was composed of a Python script that predicted PFS (anti-concordant) from PET/CT, tumor masks, and clinical data (user supplied) by accessing previously built 10-fold cross validation models in the form of. pth files. Two BASH scripts were included in the Docker image that allowed organizers to access file repositories for test data and output a .csv file of predicted PFS. While the Docker images were composed with minimal dependencies and small Python distributions, each zipped file was ∼3GB in size.

## 3 Results and Discussion

The 10-fold cross-validation results for each set for the model implementing PET/CT images, tumor segmentation masks, and clinical data are shown in **Figure 2**. Individual sets were not significantly different than ground truth PFS except for sets 6 and 7 (p<0.05). The overall mean C-index across all folds was 0.855 ± 0.060 and 0.65 ± 0.074 when considering and not considering observed events in the C-index calculation, respectively. For comparison, the equivalent model without segmentation data achieved mean C-index values of 0.841 ± 0.045 and 0.622 ± 0.067 when considering and not considering observed events in the C-index calculation, respectively. Though gains in performance were modest for the 4-channel (with segmentation) model compared to the 3-channel (without segmentation) model, these results indicate important prognostic information can further be teased out of a region of interest to improve the performance of a deep learning clinic prediction model. Upon submitting our ensemble models on external test set validation, the AVERAGE method demonstrates a C-index of 0.696 while the CONSENSUS method demonstrates a C-index of 0.698. Both these methods improve upon the analogous submissions in Task 2 of the HECKTOR Challenge, i.e. 0.689 and 0.694 for the AVERAGE and CONSENSUS models without segmentation masks, respectively. Compared to the internal validation results the increase in external validation performance for these models is likely secondary to improved generalization introduced by the ensembling approaches. Previous studies have shown that combining deep learning methods with information from segmentation masks (particularly pre-defined radiomic features) can improve outcome prediction in HNSCC [2], which is consistent with our results. In addition, manually generated tumor segmentation masks are a byproduct of physician knowledge, so it is possible the added segmentation masks allow our models to impart a degree of expert insight into predictions. An interesting future research direction would be implementing deep learning interpretability methods [12] on these models to investigate how they differ frommodels without provided segmentations.

**Fig. 2.**
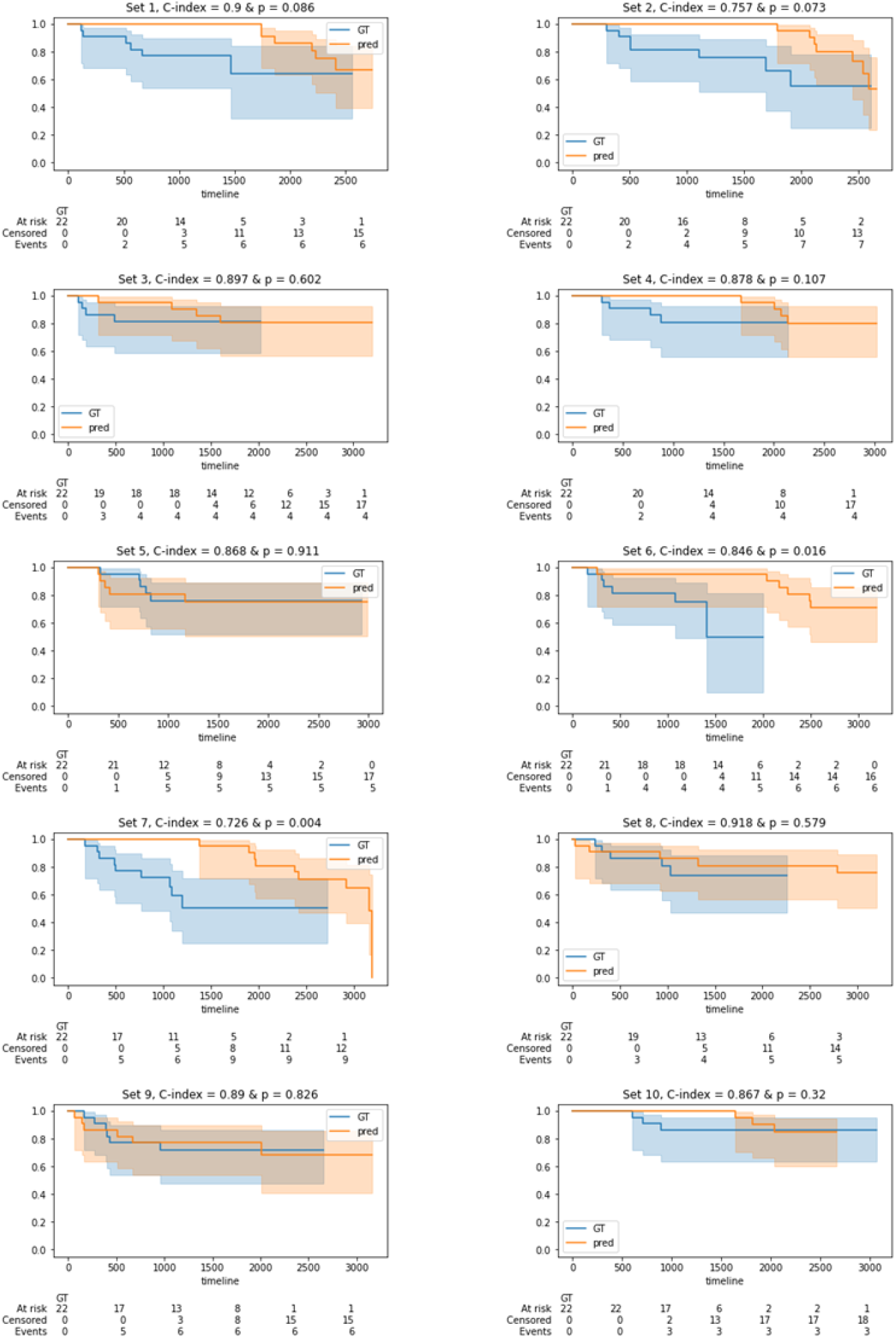
Kaplan Meier plots showing survival probabilities as a function of time in days for the ground truth (GT) PFS and the predicted PFS by the four channel (PET, CT, tumor segmentation mask, clinical) model. The C-index and the p-value of the logrank test for the GT and predicted PFS are shown above each subplot. The tables below each plot correspond to the number of patients experiencing GT PFS at each time-step.

## 4 Conclusion

Herein, we have developed and investigated the performance of a deep learning model that can utilize PET/CT images, tumor segmentation masks, and clinical data simultaneously to predict PFS in HNSCC patients. This approach is innovative since most deep learning techniques for medical outcome prediction typically only implement images and/or clinical data as model input channels. Our models achieve reasonable performance through internal-validation and external test set validation on large heterogenous datasets provided by the 2021 HECKTOR Challenge. Moreover, our results indicate subregions of interest acting as separate input channels could help add prognostic value for HNSCC prognostic deep learning models and should be investigated further.

## Data Availability

Data are available from the MICCAI HECKTOR 2021 Challenge Organizers.

## Acknowledgements

M.A.N. is supported by a National Institutes of Health (NIH) Grant (R01 DE028290-01). K.A.W. is supported by a training fellowship from The University of Texas Health Science Center at Houston Center for Clinical and Translational Sciences TL1 Program (TL1TR003169), the American Legion Auxiliary Fellowship in Cancer Research, and a NIDCR F31 fellowship (1 F31 DE031502-01). C.D.F. received funding from the National Institute for Dental and Craniofacial Research Award (1R01DE025248-01/R56DE025248) and Academic-Industrial Partnership Award (R01 DE028290), the National Science Foundation (NSF), Division of Mathematical Sciences, Joint NIH/NSF Initiative on Quantitative Approaches to Biomedical Big Data (QuBBD) Grant (NSF 1557679), the NIH Big Data to Knowledge (BD2K) Program of the National Cancer Institute (NCI) Early Stage Development of Technologies in Biomedical Computing, Informatics, and Big Data Science Award (1R01CA214825), the NCI Early Phase Clinical Trials in Imaging and Image-Guided Interventions Program (1R01CA218148), the NIH/NCI Cancer Center Support Grant (CCSG) Pilot Research Program Award from the UT MD Anderson CCSG Radiation Oncology and Cancer Imaging Program (P30CA016672), the NIH/NCI Head and Neck Specialized Programs of Research Excellence (SPORE) Developmental Research Program Award (P50 CA097007) and the National Institute of Biomedical Imaging and Bioengineering (NIBIB) Research Education Program (R25EB025787). He has received direct industry grant support, speaking honoraria and travel funding from Elekta AB.

## References

1. Wang, X., Li, B.: Deep Learning in Head and Neck Tumor Multiomics Diagnosis and Analysis: Review of the Literature. Front. Genet. 12, 42 (2021).

2. Diamant, A., Chatterjee, A., Vallières, M., Shenouda, G., Seuntjens, J.: Deep learning in head & neck cancer outcome prediction. Sci. Rep. 9, 1 –10 (2019).

3. Afshar, P., Mohammadi, A., Plataniotis, K.N., Oikonomou, A., Benali, H.: From handcrafted to deep-learning-based cancer radiomics: challenges and opportunities. IEEE Signal Process. Mag. 36, 132–160 (2019).

4. Mohamed, A.S.R., Rosenthal, D.I., Awan, M.J., Garden, A.S., Kocak-Uzel, E., Belal, A.M., El-Gowily, A.G., Phan, J., Beadle, B.M., Gunn, G.B.: Methodology for analysis and reporting patterns of failure in the Era of IMRT: head and neck cancer applications. Radiat. Oncol. 11, 1–10 (2016).

5. Paidpally, V., Chirindel, A., Lam, S., Agrawal, N., Quon, H., Subramaniam, R.M.: FDG-PET/CT imaging biomarkers in head and neck squamous cell carcinoma. Imaging Med. 4, 633 (2012).

6. Andrearczyk, V., Valentin, O., Mario, J., Vallières, M., Castelli, J., Elhalawani, H., Boughdad, S., Prior, J.O., Depeursinge, A.: Overview of the HECKTOR challenge at MICCAI 2020: Automatic Head and Neck Tumor Segmentation in PET/CT.

7. Andrearczyk, V., Oreiller, V., Vallières, M., Castelli, J., Elhalawani, H., Jreige, M., Boughdad, S., Prior, J.O., Depeursinge, A.: Automatic segmentation of head and neck tumors and nodal metastases in PET -CT scans. In: Medical Imaging with Deep Learning. pp. 33–43. PMLR (2020).

8. Iandola, F., Moskewicz, M., Karayev, S., Girshick, R., Darrell, T., Keutzer, K.: Densenet: Implementing efficient convnet descriptor pyramids. arXiv Prepr. 1404.1869. (2014).

9. Project MONAI.

10. Davidson-Pilon, C.: lifelines: survival analysis in Python. J. Open Source Softw. 4, 1317 (2019).

11. Merkel, D.: Docker: lightweight linux containers for consistent development and deployment. Linux J. 2014, 2 (2014).

12. Singh, A., Sengupta, S., Lakshminarayanan, V.: Explainable deep learning models in medical image analysis. J. Imaging. 6, 52 (2020).

